# SARS-CoV-2 Transmission Risk from sports Equipment (STRIKE)

**DOI:** 10.1101/2021.02.04.21251127

**Authors:** Thomas Edwards, Grant A Kay, Ghaith Aljayyoussi, Sophie I Owen, Andy R Harland, Nicholas S Pierce, James D F Calder, Tom E Fletcher, Emily R Adams

**Affiliations:** Department of Tropical Disease Biology, Liverpool School of Tropical Medicine, Pembroke Place, Liverpool, L3 5QA; Wolfson School of Mechanical, Manufacturing and Electrical Engineering, Loughborough University, Ashby Road, Loughborough, LE11 3TU; England and Wales Cricket Board and National Centre for Sport and Exercise Medicine, Loughborough University, LE11 3TU; Fortius Clinic, London, W1U 2EU; Department of Bioengineering, Imperial College London, London, SW7 2AZ; Department of Clinical Sciences, Liverpool School of Tropical Medicine, Pembroke Place, Liverpool, L3 5QA

## Abstract

**OBJECTIVES:** To investigate the potential of shared sporting equipment as transmission vectors of SARS-CoV-2 during the reintroduction of sports such as soccer, rugby, cricket, tennis, golf and gymnastics.

**SETTING:** Laboratory based live SARS-CoV-2 virus study

**INTERVENTIONS:** Ten different types of sporting equipment were inoculated with 40μl droplets containing clinically relevant high and low concentrations of live SARS-CoV-2 virus. Materials were then swabbed at time points relevant to sports (1, 5, 15, 30, 90 minutes). The amount of live SARS-CoV-2 recovered at each time point was enumerated using viral plaque assays, and viral decay and half-life was estimated through fitting linear models to log transformed data from each material.

**MAIN OUTCOME MEASURE:** The primary outcome measure was quantification of retrievable SARS-CoV-2 virus from each piece of equipment at pre-determined time points.

**RESULTS:** At one minute, SARS-CoV-2 virus was recovered in only seven of the ten types of equipment with the low dose inoculum, one at five minutes and none at 15 minutes. Retrievable virus dropped significantly for all materials tested using the high dose inoculum with mean recovery of virus falling to 0.74% at 1 minute, 0.39% at 15 minutes and 0.003% at 90 minutes. Viral recovery, predicted decay, and half-life varied between materials with porous surfaces limiting virus transmission.

**CONCLUSIONS:** This study shows that there is an exponential reduction in SARS-CoV-2 recoverable from a range of sports equipment after a short time period, and virus is less transferrable from materials such as a tennis ball, red cricket ball and cricket glove. Given this rapid loss of viral load and the fact that transmission requires a significant inoculum to be transferred from equipment to the mucous membranes of another individual it seems unlikely that sports equipment is a major cause for transmission of SARS-CoV-2. These findings have important policy implications in the context of the pandemic and may promote other infection control measures in sports to reduce the risk of SARS-CoV-2 transmission and urge sports equipment manufacturers to identify surfaces that may or may not be likely to retain transferable virus.

WHAT IS ALREADY KNOWN ON THIS TOPIC

- Transmission of SARS-CoV-2 between individuals playing sport may be via respiratory droplets when in close proximity to an infected person.
- SARS-CoV-2 remains viable on a variety of surfaces resulting in recommendations to reduce the sharing of sports equipment such as tennis balls when sports were re-opened.

WHAT THIS STUDY ADDS

- The recoverable SARS-CoV-2 viral load reduces exponentially with mean viral load of all materials less than 1% of the original inoculum after 1 minute.
- The type of material has a significant effect on SARS-CoV-2 transfer, with less virus transferred from porous materials such as bovine leather or nylon woven cloth.
- Policies on infection control measures in sport may be better directed towards areas other than reducing the sharing of sports equipment.
- Sports equipment manufacturers may consider using materials that absorb or retain virus as a way of reducing viral transmission from sports equipment.

## Introduction

Public health interventions to control the COVID-19 pandemic in the UK has necessitated restrictions in social mixing, with both amateur and professional sports either prohibited, or allowed with considerable infection control measures in place.^1^ It has been estimated up to 44% of transmission occurs prior to symptom onset, when viral loads are highest,^2^ meaning people are unaware of the infection and will continue in their daily activities, including sports participation. All team sport in the UK was postponed and subsequently cancelled from mid-March 2020 and although a staged return to elite sport was enabled from April,^3^ there was concern that participation in community sport could risk disease transmission. Limited return in team sport with strict hygiene measures at sports grounds and rule changes for some contact sports were imposed.

The major risk of SARS-CoV-2 transmission during team sports is likely direct player to player transmission via respiratory droplets in close proximity to an infected person,^4^ either during play or socially before and after the game. During non-contact sports such as cricket and soccer the risk of transmission is considered very low because only fleeting incursions of social distancing are seen.^5^ Few amateur sporting events have been linked to SARS-CoV-2 transmission. A Danish study on professional football reported that during a 90 minute match the average time any player spent within 1.5m of another was 87.8 seconds,^12^ and a study of the return of competitive football in Germany concluded that training and matches may be carried out safely during the SARS-CoV-2 pandemic.^13^ Although there is uncertainty about the role of fomites in SARS-CoV-2 transmission,^6^ equipment which commonly shared in sports is potentially an important route of transmission. As a result, when team sports were re-introduced, so too were recommendations to reduce the sharing of sports equipment such as tennis balls. Whilst the viability of virus on a variety of surfaces has been demonstrated, transmission requires the deposition of virus onto a surface, then the transfer of enough virus to cause an infection from that surface to the mucus membranes of another person. Viral shedding into the environment has been demonstrated during SARS-CoV-2 infections, for example the rooms of hospitalised SARS-CoV-2 patients can be heavily contaminated with SARS-CoV-2, including frequently touched surfaces such as sinks and door handles.^8^ This is thought to be from the spread of respiratory droplets via breathing, sneezing and coughing,^9^ and the transfer of SARS-CoV-2 to objects from the hands of patients has been documented.^10^ The minimum infectious dose of SARS-CoV-2 is currently unknown,^7^ which makes quantifying the required viral load for transmission difficult.

It has been hypothesised that because physical sports lead to increased respiration, deep exhalation is likely to increase the expulsion of droplets or aerosols containing infectious particles which may contaminate sports equipment. Additionally, spitting is common in some team sports such as football and rugby and saliva has been used to shine cricket balls. Secretions from the upper respiratory tract have the potential to carry high viral loads,^11^ which could then be transferred onto materials such as clothing or balls. Studies on viral stability on sporting equipment are limited. Pelisser et al reported that inactivated SARS-CoV-2 may be detected by RT-qPCR from the surfaces of cricket balls up to one hour past the inoculation,^14^ although viral viability could not be determined using this approach. Determining the potential for SARS-CoV-2 transmission whilst sharing sports equipment is crucial when producing guidelines to mitigate risks for return of community and elite team sports. This study [SARS-CoV-2 Transmission Risk from sports Equipment (STRIKE)] aimed to quantify the recoverable live virus from droplets of viable SARS-CoV-2 suspensions deposited onto a variety of sporting equipment, over a time frame relevant to sporting activity.

## Methods

### Materials

Sporting materials were selected from the materials collection at the Sports technology Institute at Loughborough University (Table 1). All materials were unused prior to testing. Materials frequently shared between participants were prioritised due to a greater potential as transmission vectors. Materials were cut into 2cm diameter disks using a metal hole punch. The red cricket ball was not cut into disks, and instead used whole, due to difficulties retaining surface integrity during removal. Materials were not sterilised prior to inoculation, so as not to affect the surface coatings, and antibiotics in the media were relied upon to avoid contamination during cell culture. Steel disks were purchased (Lasermaster, UK) for use as a control surface.

**Table 1.** Equipment and materials used in the study. * indicates standards available to certify such products, however It is not known whether product was subject to standard testing.

| **Equipment type** | **Standard** | **Surface Material** |
| --- | --- | --- |
| Cricket glove (palm) | BS 6183-4: 2001* | Calf leather |
| Control | Stainless steel |  |
| Football | FIFA Quality Pro. Law 2: IFAB Laws of the Game | Thermoplastic Polyurethane (TPU) |
| Golf ball | Part 4, The Equipment Rules, USGA. Named on list of conforming balls, USGA, R&A | Surlyn™ ionomer resin |
| Gym pit foam | n/a | Polyurethane (PU) foam (open cell) |
| Horse saddle | n/a | Polyurethane (PU) |
| Red Cricket ball (unused) | BS 5993:1994* Law 4, The Laws of Cricket, MCC | Bovine leather burnished with synthetic grease |
| Rugby ball | Law 2: Ball, World Rugby Laws | Rubberised polyester (PES) |
| Tennis Ball | ITF Approved, Rule 3, ITF Rules of Tennis | Raised Wool/Nylon woven cloth |
| White Cricket ball (unused) | BS 5993:1994* Law 4, The Laws of Cricket, MCC | Bovine leather with nitrocellulose coating |

### Cell culture

Cultures of VERO E6 cells (C1008; African green monkey kidney cells, European Collection of Authenticated Cell Cultures 85020206) were maintained in T75 cell culture flasks (Corning, US) in Dulbecco’s Modified Eagles Medium (DMEM) supplemented with 4.5g/L glucose and L-Glutamine (Lonza, US), 10% foetal bovine serum (Sigma, US) and 50 units per ml of penicillin/streptomycin (Gibco, US), at 37.5°C + 5% CO2. Cells for plaque assays were detached from the monolayer using 2ml 1x trypsin-EDTA (Sigma, US) and 500μl was seeded into 24 well microtitre plates (Corning, US) at a density of 250,000 cells/ml. Plates were incubated for 24 hours at 37.5°C + 5% CO2 and used for downstream plaque assays if judged to be >95% confluent by microscopy.

### Material inoculation

Materials were inoculated with a 40μl droplet of DMEM containing a high (1.2×10^6^ plaque-forming units (PFU)/ml) or low (1.2×10^4^ PFU/ml) concentration of quantified live SARS-CoV-2 virus (isolate REMRQ0001/Human/2020/Liverpool). Inoculum concentrations were chosen as representing the upper and lower quartile of viral loads in symptomatic patients.^15^ All work with live virus took place under BSL3 conditions in a Class 2 biological safety cabinet. Materials were inoculated on the outward facing surface, and care was taken to ensure the inoculum did not run off the material during the inoculation. Triplicate pieces of each material were inoculated for the following time points: 1, 5, 15, 30, 90 minutes. At each time point the materials were swabbed using dry cotton swabs (Copan, Italy), and added to 400μl of DMEM. A standardised swabbing technique was employed for each sample to reduce variation, with the swab being dragged upwards for two seconds and sideways for two seconds. The tubes containing the swabs were vortexed for 5 seconds, and then serially diluted ten-fold for three dilutions, in DMEM.

### Viral plaque assays

Viral plaque assays were carried out using VERO E6 cells in 24 well microtitre plates. Media was aspirated from the microtitre plates, and 40μl of media from the swabs and further dilutions were added to triplicate wells with 160μl of DMEM 2% FBS. Microtitre plates were incubated for 1 hour at 37.5°C + 5% CO2 to ensure viral infection. Plates were then removed from the incubator and overlaid with a 1.1% suspension of cellulose (Sigma, UK) in DMEM 2% FBS, and incubated again under the same conditions for 72 hours. Plates were then removed from the incubation, fixed with 100% formaldehyde for one hour, and stained with 1ml/well of 0.25% crystal violet solution. After staining for 1 minute, plates were gently washed with water, then air dried for >3 hours. Viral plaques were then visually counted for each well.

### Outcomes

All individual plaque counts for each material/time point swab were used to analyse the viral recovery from each material. Readings below the limit of quantitation (BLQ) were assumed to be equal to the BLQ levels / 2. To characterise SARS-CoV-2 retained on different materials, a dynamic approach was used to measure the time-reduction course of virus. This was achieved by estimating viral decay half-life through linear models to log transformed PFU data on each material. The model assumed a single-phase decay profile on all materials over time. Each material was assumed to have a different intercept and slope as the materials varied widely in the observed initial levels of virus at the first minute.

### Statistical analysis

Linear fitting was achieved through the lm function in R. After generating a slope and intercept for each material, simulations were performed based on the estimates for each of these parameters and their covariance using R. 500 simulations were performed for each material and their 5-95 percentiles were plotted against the observed data. To estimate the overall exposure to virus over time for each material, the area under the curve (AUC) of the time-virus simulated profile was estimated for each of the 500 simulations and compared as the primary measure describing the overall exposure of each material to the virus over time. Simulated AUCs for each material were compared statistically using non-parameteric pairwise comparisons using Wilcoxon rank sum test. Materials were ranked based on AUC in comparison to the control material on all reported graphs. Simulated half-life and AUC data are presented as box and whiskers plots displaying the 5-95 percentiles of the 500 simulated profiles for each material.

### Patient and Public Involvement statement

As this was solely a laboratory study we did not involve any patients or members of the public in the study design.

## Results

At the one-minute time point SARS-CoV-2 was only detected in 7/10 materials when using the low inoculum dose (recovered virus ranging from 10 to 40 virions) (Fig. 1). At five minutes virus was detected on the horse saddle alone (20 virions) and no virus was detected on any material at 15 minutes. The model-predicted decay for the low inoculum is shown in Fig. 2 and the AUC and predicted half-life for the low inoculum were lower than that of the high inoculum (Fig. 3).

**Figure 1.**
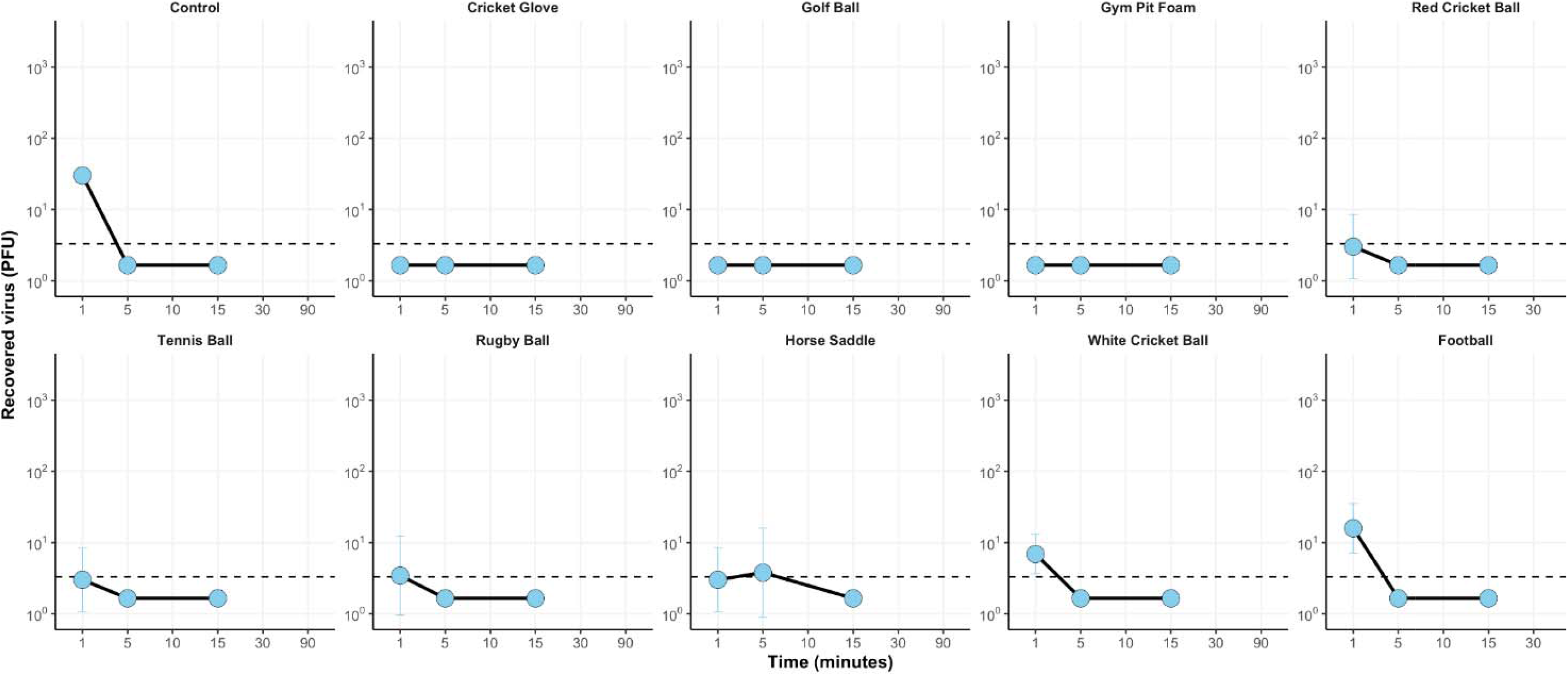
Recovered virus from all materials inoculated with 5.4×10^2^PFU (low inoculum) across the 90 minute sampling time. Points represent the geometric mean of three replicates and Error bars indicate the geometric standard deviations.

**Figure 2.**
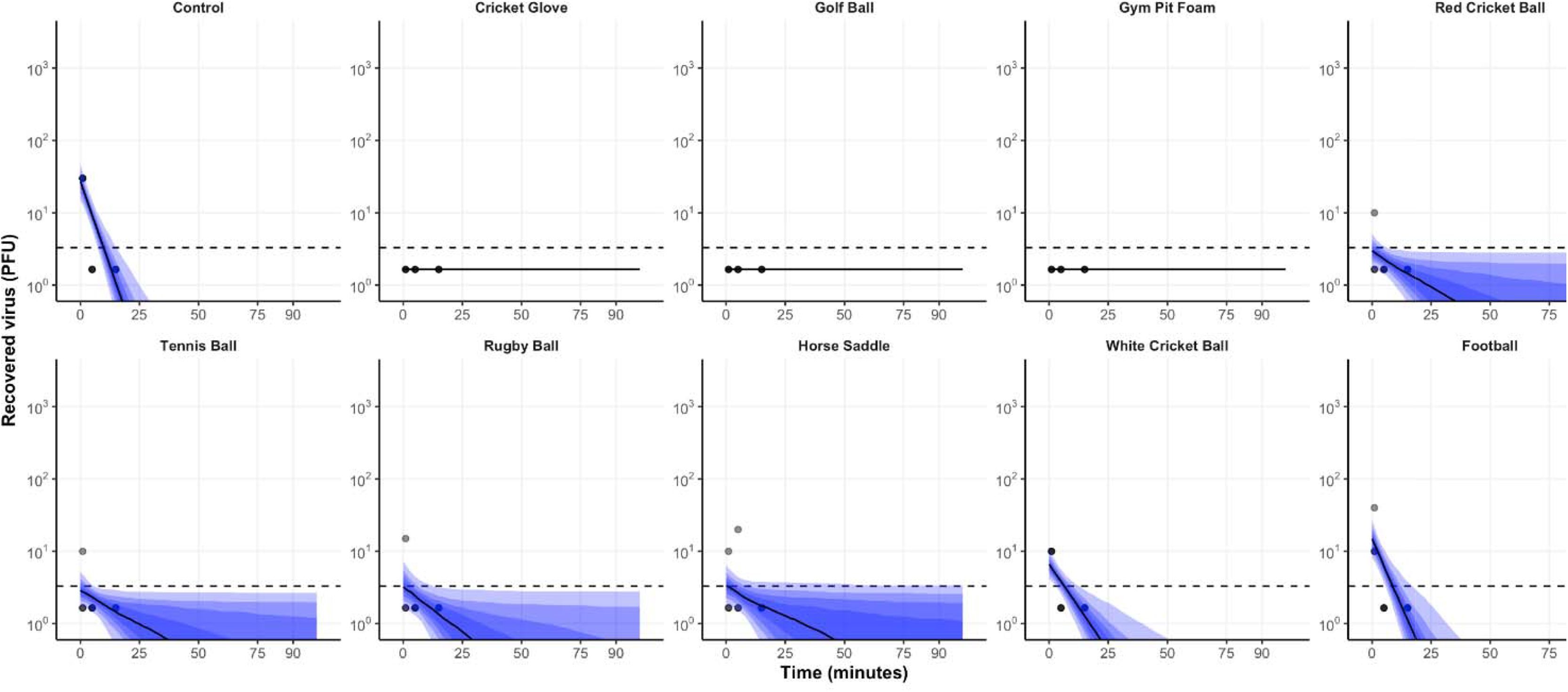
The predicted median decay of viral titres (solid black line) with 5-95 percentiles (shaded red areas) overlaid with observed PFU (grey circles) from all materials inoculated with 5.4×10^2^ PFU (low inoculum) using a linear regression model on log-transformed data.

**Figure 3.**
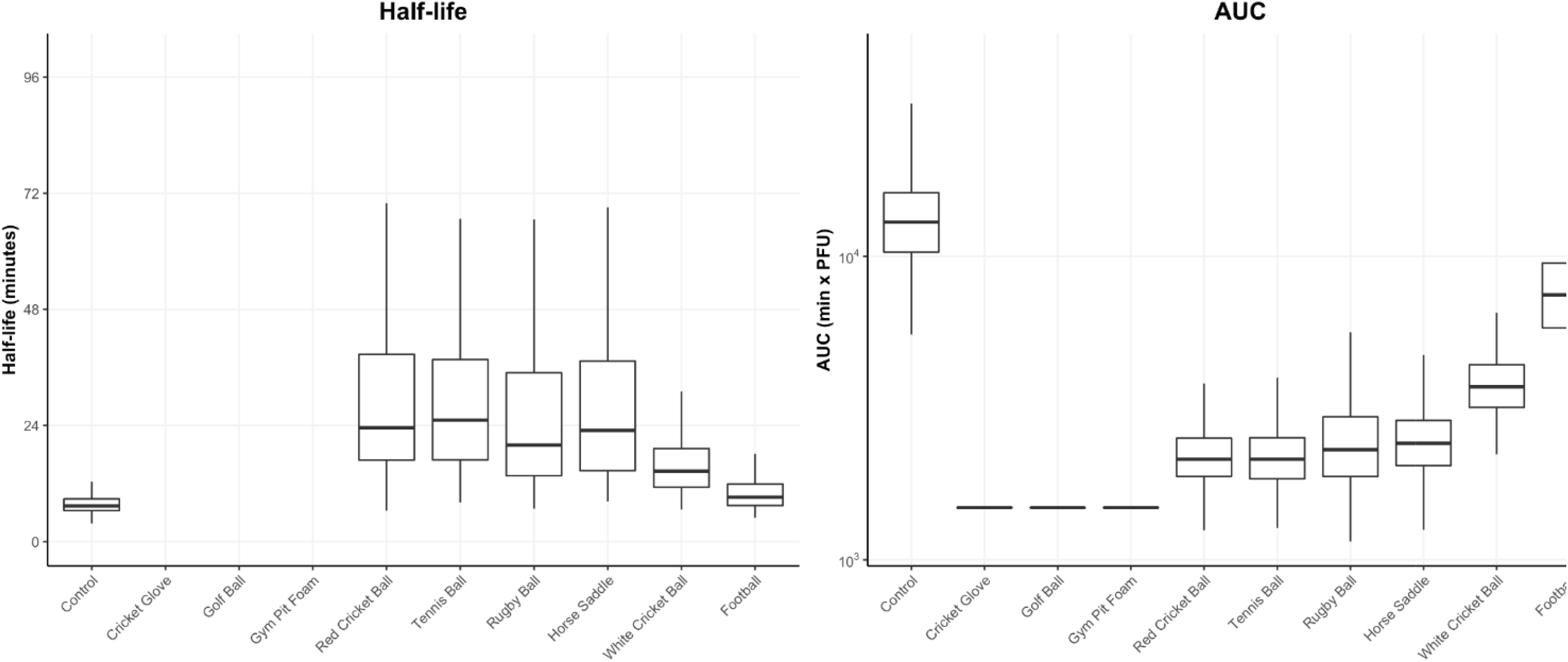
Box and whiskers plots representing the Half-life and area under the curve (AUC) distribution of 500 generated profiles for each material inoculated with SARS-CoV-2 at 5.4×10^2^ PFU (low inoculum).

For the high inoculum dose, virus was recoverable from every material except the cricket glove at the one-minute time point (Fig. 4) but the viral titre reduced from 5.4×10^4^ to an average of 3.7×10^2^ (range of 1×10^3^ to 0) virions. The highest recovery was obtained from the rugby ball, steel control disks and horse saddle with the lowest viral titres retrieved were the from absorbent materials such as the cricket glove, red cricket ball and tennis ball. Viral recovery reduced over time for all materials tested and no virus could be retrieved at 90 minutes, except for the horse saddle and rugby ball, although viral levels had reduced to 2 and 12 virions, respectively. The mean recovery reduced significantly at the one, five, 15, and 30-minute time points for all materials (P=0.0137, 0.0185, 0.0174, and 0.0117, respectively). The mean recovery of virus fell across all materials to 0.74% at one minute, 0.39% at 15 minutes and 0.003% at 90 minutes (Fig. 5).

**Figure 4.**
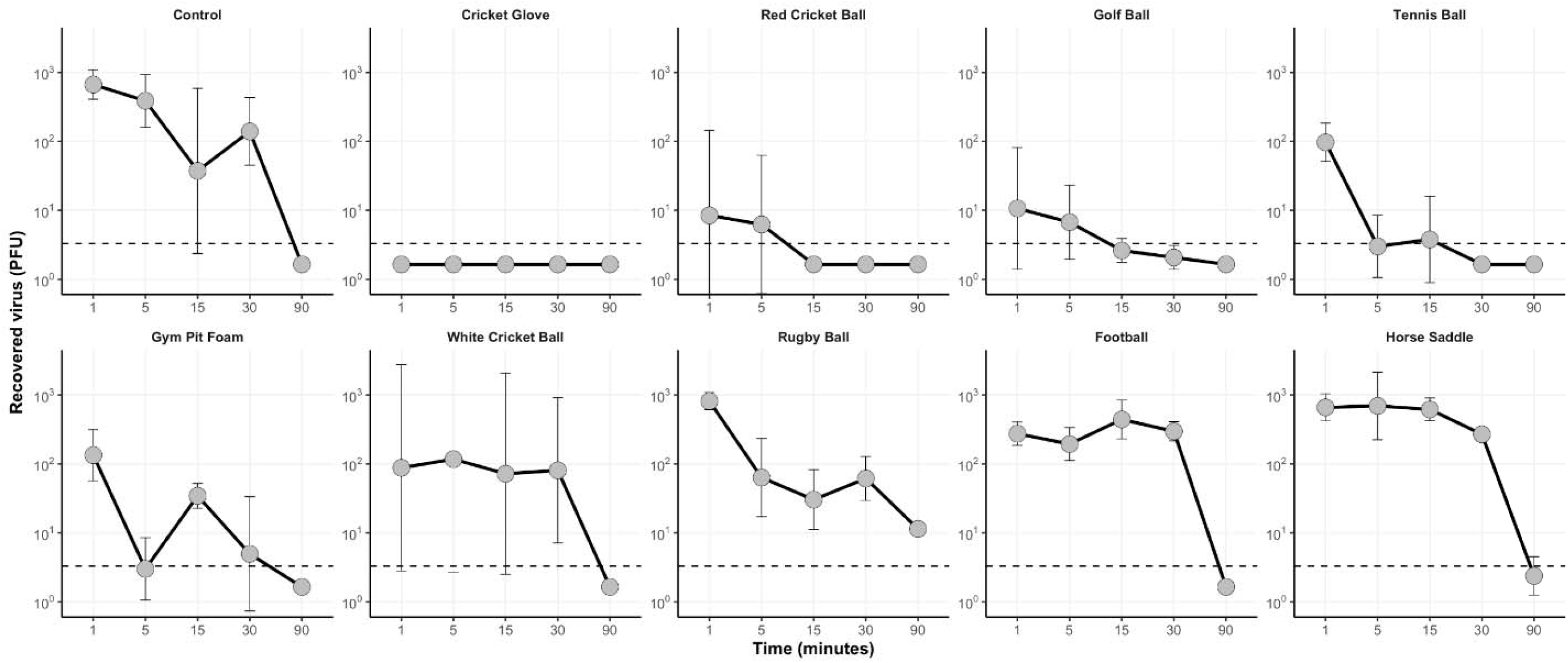
Recovered virus from all materials inoculated with 5.4×10^4^ PFU (high inoculum) across the 90-minute sampling time. Points represent the geometric mean of three replicates and Error bars indicate the geometric standard deviations.

**Figure 5.**
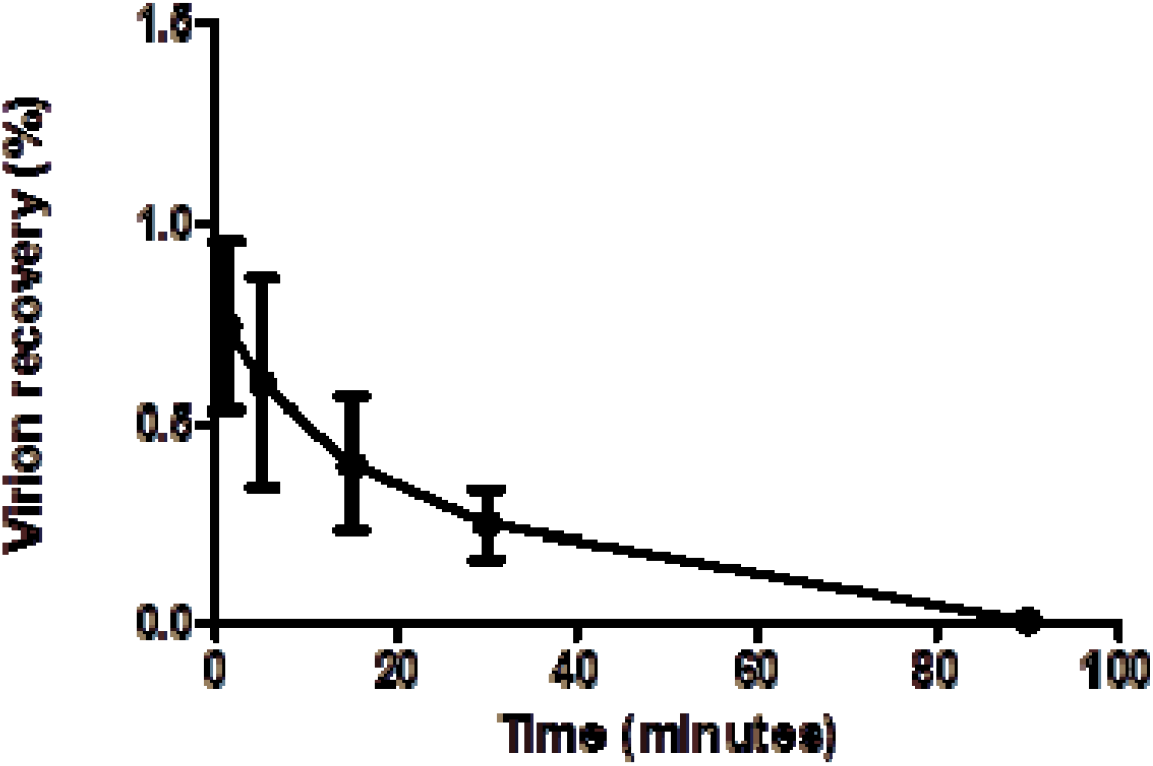
Proportion of virions recovered from high inoculum of 5.4×10^4^ from all materials at each time point. Error bars represent the standard error of the mean.

Exponential viral decay was predicted from the linear regression models for all surfaces tested, indicated by a straight line of decay on the log^10^ viral PFU scale (Fig. 6). Data from the white cricket ball was more varied, with subsequent wider ranges in possible decay rates. The estimated mean half-life of deposited virus after the one minute time point ranges from 24 minutes (control) to 60 minutes (golf ball) (Fig. 7). The area under the curve (AUC) analysis takes into account the initial decrease in recoverable virus, and ranks the surfaces in terms of transmissibility of SARS-CoV-2, from porous materials such as the cricket ball to less porous materials such as the horse saddle (Fig. 7). Analyses using Wilcox non-parametric test showed that AUCs simulated for different materials are statistically different with p<0.001 with the horse saddle containing the highest amount of virus over time and the cricket glove the lowest. The model-predicted decay for the low inoculum is shown in Fig 2 and the AUC and predicted half-life for the low inoculum were lower than that of the high inoculum (Fig.3).

**Figure 6.**
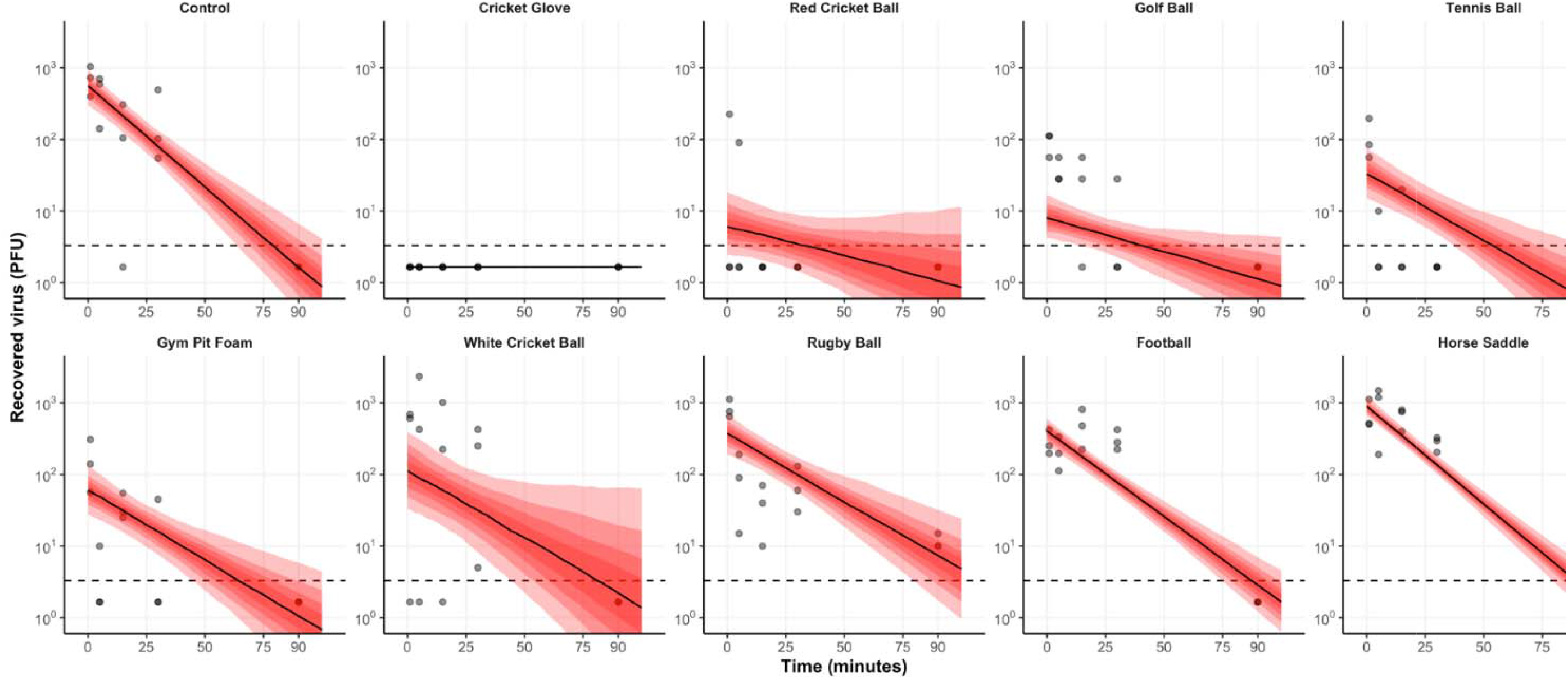
The predicted median decay of viral titres (solid black line) with 5-95 percentiles (shaded red areas) overlaid with observed PFU (grey circles) from all materials inoculated with 5.4×10^4^ PFU (high inoculum) using a linear regression model on log-transformed data.

**Figure 7.**
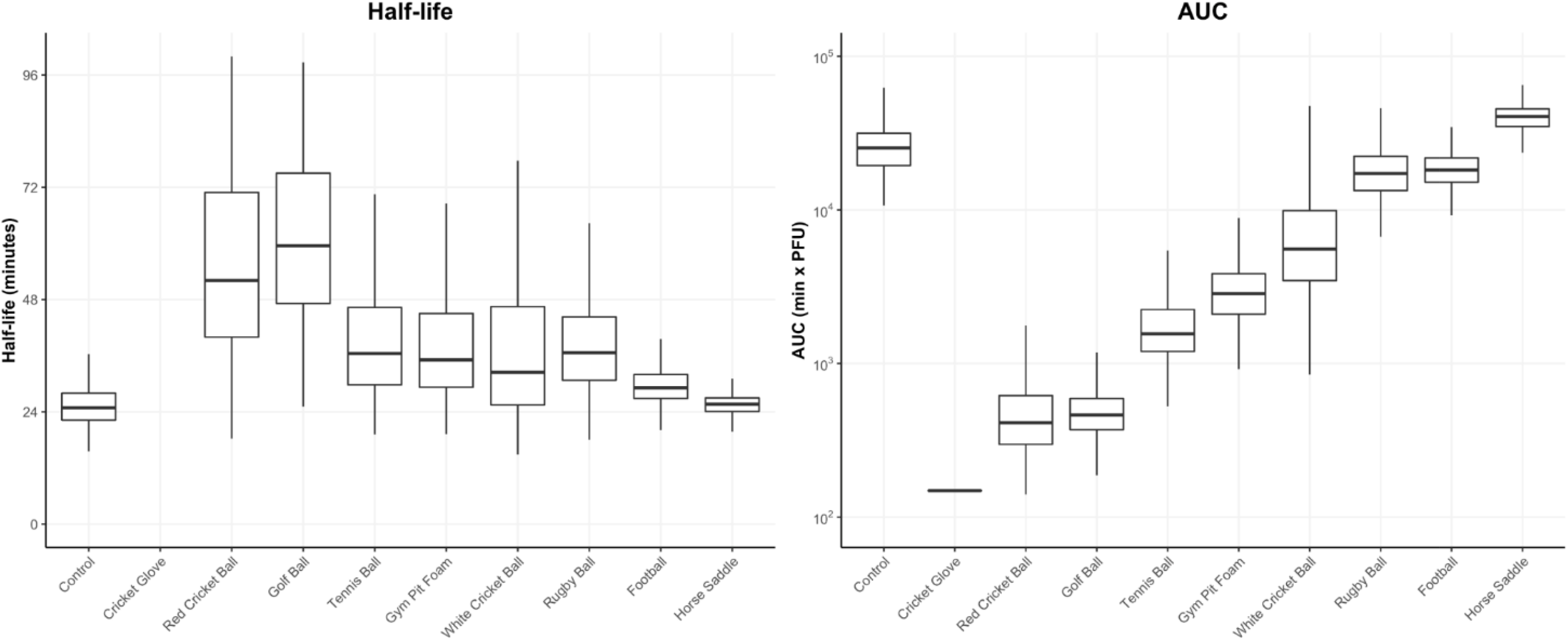
Box and whiskers plots representing the Half-life and area under the curve (AUC) distribution of 500 generated profiles for each material inoculated with SARS-CoV-2 at 5.4×10^4^ PFU (high inoculum)

## Discussion

This live virus laboratory based study demonstrates an exponential reduction in detectable SARS-CoV-2 virions for all inoculated sports equipment over a very short time period. The low inoculum (representing droplets from the lower quartile of viral loads in symptomatic patients)^15^ could only be detected on the horse saddle at five minutes and no virus could be detected on any material at 15 minutes. 0.74% of virus was recoverable at one minute in the high inoculum (representing droplets from the higher quartile of viral loads), 0.39% at 15 minutes and just 0.003% at 90 minutes. This indicates that transfer of sufficient virus from fomites is unlikely from individuals with lower viral loads. As the viral inoculum dried over time, less virus was recoverable, matching previous studies on inanimate surfaces.^16^

The contribution of fomites to the transmission of SARS-CoV-2 is controversial.^19^ Wilson *et al*. ^20^ used a Monte Carlo simulation to perform a quantitative microbial assessment of the risk of infection from fomites, and found a lower than 1/10,000 infection risk from surfaces infected with a range of 1 to 10,000 genome copies/cm, supporting the potentially limited role of fomites in SARS-CoV-2 transmission. In addition, it should be noted that in certain sports, equipment such as balls may come in contact with crowds where unknown viral load may be encountered. Participants in sports are likely to be asymptomatic, however viral loads in this sub group appears to be similar to those in symptomatic patients.^17^ A South Korean cohort study of 303 non-hospitalised SARS-CoV-2 positive patients (193 symptomatic/110 asymptomatic) found no significant difference in RT-qPCR Ct values between the groups.^18^ Quantification of the viral load that may be transferred from an individual with SARS-CoV-2 infection onto sports equipment has not been evaluated but this study used previously reported concentrations seen in respiratory tract secretions.

### The effect of material composition on SARS-CoV-2 transfer

We found that viral recovery was reduced by absorbent materials such as leather (red cricket ball and cricket glove) and polyurethane foam (gym mat foam). Despite the white and red cricket ball surfaces both being composed of bovine leather, the different coatings used to finish the surfaces (synthetic grease on the red ball, nitrocellulose on the white ball) had a noticeable effect on viral recovery, with the red ball having a lower level. These properties were observed in unused cricket balls. It is expected that with use cricket balls will lose the coatings and may become more porous. Previous studies have shown that viruses such as avian influenza have shorter recovery times from porous materials or open-cell foam structures,^21^ presumably as viral particles are trapped inside and are not easily transferred during contact. The physiochemical interactions between the viral capsid and the material is also likely to impact the viral viability and transfer from the material,^22^ and therefore the hydrophobicity and electrostatic properties of polymer surfaces may be important in their role as fomites. The observation that porous materials result in reduced viral recovery and transmission risk can be used to prioritise materials for within-game cleaning or swapping, and focus cleaning efforts to reduce their effect on sporting events.

### Limitations

All experiments were carried under a single temperature and humidity, two parameters known to affect SARS-CoV-2 viability.^26^ Sports are played under different conditions due to seasonality and whether they take place in or outdoors. A more accurate assessment would include these variables. The minimum infectious dose of SARS-CoV-2 remains unknown,^7^ and this makes it difficult to extrapolate the amount of virus on a surface necessary for transmission. The prospect of future human challenge models may provide this data.^27^ Our methodology employed a dry swab to retrieve the virus from the surfaces, in order to best replicate the transfer onto a player’s body or clothing. Higher viral recovery, and possibly less variation between replicates, would have been achieved by directly adding media to absorb virus^23^ or by using a wet swab.^24^ However, this would not replicate the real-world conditions that the experiments were designed to assess. The recovery rate of dry swabs varies, but has been estimated as 32-38% for recovering MS2 phage from steel surfaces, depending on the elution media.^25^ Therefore, more virus is likely to be present on the materials than our results may infer.

This was a laboratory study, and further in-game behavioural studies are required to show frequency of potential transmission events to quantify risk. In practice many items of sports equipment are not routinely handed from person to person but instead rub against or collide with implements, other parts of the body, the ground and other sports infrastructure, presumably further reducing viral load. This is illustrated by a study showing SARS-CoV-2 RNA was not detectable from inoculated cricket balls after wiping, rolling, or bouncing on the floor.^14^

### Conclusions

This study demonstrates the rapid decay in transmissible SARS-CoV-2 virus on several types of sports equipment and given the uncertainty of the role of fomites in the transmission of virus it is likely that close contact with other players either during play or pre/post-match travel and socialising is more important as a mode of spreading the virus. This has implications for policymakers introducing control measures during the reopening of sports. The differences in transfer seen between types of sports equipment may also direct the engineering of materials that retain and absorb virus, as opposed to hydrophobic materials where viral transfer is greater.

## Data Availability

Data is available from the authors upon request

## Funding

The study was funded by philanthropic donations and contributions from the UK National Governing Bodies of Sport. The funders had no input into the design, interpretation or reporting of the study.

## Author contributions

JDFC, TEF, and ERA conceptualised the study. TE, GAK, SIO, ARH, NSP, JDFC, TEF and ERA contributed to the experimental design and methodology. TE, GAK, and SIO carried out laboratory experiments. TE and GA contributed to data curation and analysis. TE, ARH, NSP, JDFC and ERA wrote and edited the first draft of the manuscript. All authors reviewed and edited the final manuscript.

## Competing Interests

NSP is employed in part by the England and Wales Cricket Board. All other authors declare no competing interests.

## Notes

### Author Declarations

This was a laboratory study and required no ethical approval

